# Comparative Analysis of Major Incident Triage Tools in Children – a UK population-based analysis

**DOI:** 10.1101/2021.06.29.21259604

**Authors:** J Vassallo, S. Chernbumroong, N. Malik, Y. Xu, D. Keene, G.V. Gkoutos, MD. Lyttle, J.E. Smith, in collaboration with PERUKI (Paediatric Emergency Research in the UK and Ireland)

**Affiliations:** Institute of Naval Medicine, Gosport, United Kingdom; Academic Department of Military Emergency Medicine, Royal Centre for Defence Medicine, Birmingham, United Kingdom; NIHR Surgical Reconstruction and Microbiological Research Centre (SRMRC), Heritage Building, Queen Elizabeth Hospital, Mindelsohn Way, Edgbaston, Birmingham, B15 2TH, UK; Centre for Computational Biology, Institute of Cancer and Genomic Sciences, University of Birmingham, Birmingham, B15 2TT, UK; University Hospitals Birmingham, Mindelsohn Way, Edgbaston, Birmingham, B15 2WB, UK; Institute of Translational Medicine, University Hospitals Birmingham NHS Foundation Trust, Birmingham, B15 2TT, UK; MRC Health Data Research UK (HDR UK); Emergency Department, Bristol Royal Hospital for Children, Bristol, UK; Faculty of Health and Applied Sciences, University of the West of England, Bristol, UK; Emergency Department, University Hospitals Plymouth NHS Trust, Plymouth, United Kingdom

**Author notes:** **Corresponding author** Dr J Vassallo. Institute of Naval Medicine, Alverstoke, Gosport, PO12 2DL, UK. **Funding** This study is funded by the National Institute for Health Research (NIHR) Surgical Reconstruction and Microbiology Research Centre. GVG also acknowledges support from the MRC Health Data Research UK (HDRUK/CFC/01). Additionally, the lead author received a RCEM Young Investigator grant to support Open Access publication fees for the study. **Author Contributions** JV, ML, JES designed the study. JV, SC, NM, YX verified the underlying data. JV and SC conducted the analysis. All authors contributed to data interpretation. JV wrote the initial draft of the manuscript. All authors contributed with critical revisions to the manuscript. JV takes responsibility for the manuscript as a whole.

**Keywords:** Triage, paediatrics, major incidents, life-saving interventions

## Abstract

**Introduction:** Triage is a key principle in the effective management of major incidents. There is currently a paucity of evidence to guide the triage of children. The aim of this study was to perform a comparative analysis of nine adult and paediatric triage tools, including the novel ‘Sheffield Paediatric Triage Tool’ (SPTT), assessing their ability in identifying patients needing life-saving interventions (LSI).

**Methods:** A ten-year retrospective database review of TARN data for paediatric patients (<16years) was performed. Primary outcome was identification of patients receiving one or more LSIs from a previously defined list. Secondary outcomes included mortality and prediction of ISS>15. Primary analysis was conducted on patients with complete pre-hospital physiological data with planned secondary analyses using first recorded data. Performance characteristics were evaluated using sensitivity, specificity, under and over-triage.

**Results:** 15133 patients met TARN inclusion criteria. 4962 (32.8%) had complete pre-hospital physiological data and 8255 (54.5%) had complete first recorded physiological data. Male patients predominated (69.5%), sustaining blunt trauma (95.4%) with a median ISS of 9. 875 patients (17.6%) received at least one LSI.

The SPTT demonstrated the greatest sensitivity of all triage tools at identifying need for LSI (92.2%) but was associated with the highest rate of over-triage (75.0%). Both the PTT (sensitivity 34.1%) and JumpSTART (sensitivity 45.0%) performed less well at identifying LSI. By contrast, the adult MPTT-24 triage tool had the second highest sensitivity (80.8%) with tolerable rates of over-triage (70.2%).

**Conclusion:** The SPTT and MPTT-24 outperform existing paediatric triage tools at identifying those patients requiring LSIs. This may necessitate a change in recommended practice. Further work is needed to determine the optimum method of paediatric major incident triage, but consideration should be given to simplifying major incident triage by the use of one generic tool (the MPTT-24) for adults and children.

**What this paper adds?:** *Section 1: What is already known on this subject?:* Triage is a key principle in the effective management of major incidents. There is currently a paucity of evidence surrounding the use of existing paediatric major incident tools. In the UK, two methods of paediatric major incident triage exist, the Paediatric Triage Tape and the JumpSTART method. In previous studies they have demonstrated less than 50% sensitivity at identifying children in need of life-saving interventions. This study performed a comparative analysis on a UK paediatric trauma registry population and included a newly derived triage tool, the SPTT.

*Section 2: What this study adds:* The PTT and JumpSTART perform poorly (<45% sensitivity) in this paediatric trauma registry population. The SPTT and the existing adult triage tool the MPTT-24 outperform all methods. Consideration should be given to simplifying major incident triage by the use of a single generic tool for both adults and children.

## Introduction

Major incidents occur worldwide on a regular basis, when existing resources are outstripped due to the number, type, severity or location of casualties, necessitating additional support.^1^ Triage is a key principle in effectively managing major incidents, whereby patients are prioritised on the basis of their clinical acuity, typically using a simple physiological assessment as part of a triage tool.^1^ Whilst existing triage tools have been derived and validated using mortality and injury severity as outcomes of interest, neither of these reflect the acuity of the patient or the need for a life-saving intervention.^2-5^

In the UK, two paediatric major incident triage tools exist; the Paediatric Triage Tape (PTT^6^; for pre-hospital use in patients under 12 years), and JumpSTART^7^ (for in-hospital use in patients under 9 years) Both are paediatric adaptations of adult triage tools (MIMMS Triage Sieve and START respectively^1,8^) and utilise a step-wise approach to triage; the PTT^6^ uses physiological variables based on the child’s length as a proportionate surrogate for age, and JumpSTART^7^ utilises a single respiratory rate threshold. Within the UK, one further alternative method of paediatric triage has been proposed. This is the Sheffield Paediatric Triage Tool (SPTT), a paediatric adaptation of the adult MPTT-24 (supplementary figure 1). The SPTT has undergone practical testing within a simulated paediatric major incident, where it correctly triaged all patients, thereby representing a potentially viable alternative to the PTT and JumpSTART (C.O’Connell, personal communication, January 25, 2020).

Extensive research has been conducted in adult major incident triage, leading to the development and implementation of the Modified Physiological Triage Tool-24 (MPTT-24)^9^ into both UK military and civilian in-hospital practice (NHS Clinical Guidelines for Major Incidents).^10^ By contrast, there is a paucity of evidence surrounding paediatric major incident triage tools. Limited studies have evaluated the performance of existing triage tools in identifying the need for life-saving interventions, and these have demonstrated poor performance of existing UK methods^2,11^ Perhaps the greatest challenge in designing a fit for purpose paediatric major incident tool is the determination of appropriate physiological thresholds denoting the need for intervention. Normal ranges are wide, and change with age, leading to potential for confusion in those performing triage in this high stakes, high stress event.^6,12^

Ideally any study examining the performance of triage tools should be tested in the environment in which they are to be used, (i.e., within the major incident context). However due to the unpredictability of major incidents, and associated ethical implications, the feasibility of such an assessment is very low. As a result, trauma registries, which contain high numbers of seriously injured patients, are often used as a proxy to examine the performance accuracy of such triage tools.^2,13-15^

The aim of this study was to compare the performance of the MPTT-24 and its paediatric derivative, the SPTT with existing adult and paediatric triage tools using the UK Trauma Audit and Research Network (TARN) database. The primary outcome was their performance accuracy in identifying paediatric patients in need of a life-saving intervention (Priority One patients).^16^

## Method

A retrospective database review was undertaken using the TARN database for a ten year period (1 January 2008 to 31 December 2017). All paediatric (age < 16 years) trauma patients, meeting TARN inclusion criteria (Supplementary File 1), were considered eligible.

TARN is the largest trauma registry in Europe, and collects data on all patients sustaining moderate and severe trauma admitted to trauma-receiving hospitals in England and Wales. Patients are included if they fulfil one of the following criteria: admission to a critical care area; transfer for specialist care; death; or hospital admission for >3 nights.^14^ Information is submitted electronically by trained hospital co-ordinators and captures data from injury through to discharge, including interventions. Patients declared dead at scene and not conveyed to hospital are not recorded; these were therefore not included in our analyses. Owing to the nature of the TARN registry and its inclusion criteria, patients were assumed to be non-ambulant. Patients receiving one or more life-saving interventions based on a previously defined list^16^ (with adaptations for paediatric fluid resuscitation in keeping with Advanced Paediatric Life Support^12^) were considered to be Priority One (supplementary table 1).

Primary analysis for all outcomes of interest was conducted on patients with complete pre-hospital physiological data. A planned secondary analysis was conducted using first recorded physiological data, whether performed in the pre-hospital or emergency department (ED) setting. Patients were categorised as Priority One, or Not Priority One, using available paediatric (PTT^6^, JumpSTART^7^ and SPTT) and adult (MPTT-24 (including airway opening manoeuvre)^9,17^, NASMeD Sieve^18^, MSTART^19^, Careflight^14^, MIMMS Triage Sieve^1^, RAMP^20^) triage tools. In keeping with the design of the JumpSTART and PTT, once a patient reached a pre-defined age (8 and 12 years respectively), they were then triaged by the corresponding adult triage tool (START and MIMMS Triage Sieve respectively^1,18^). A comparison of the different triage tools is shown in Table 1. Assumptions made for the categorisation of triage tools is described in supplementary file 2.

**Table 1:** Triage Tool Comparison.

| Tool | Tool components |  |  |  |  |  |  |
| --- | --- | --- | --- | --- | --- | --- | --- |
|  | 1<sup>st</sup> step | 2<sup>nd</sup> step | 3<sup>rd</sup> step | 4<sup>th</sup> step | 5th step | 6<sup>th</sup> step | 7<sup>th</sup> step |
| Paediatric Triage Tape (PTT)<sup>6</sup> | <10kg:<br>Alert & moving all limbs<br>11-18kg:<br>Alert & moving all limbs or<br>Walking<br>>19kg:<br>Walking | Breathing<br>(Open Airway if<br>required) | Respiratory Rate:<br><10kg: <20 or > 50<br>11-18kg: <15 or >45<br>>19kg: <10 or > 30 | Capillary refill < 2 sec | Heart Rate:<br><10kg: <90 or > 180<br>11-18kg: <80 or > 160<br>>19kg: <70 or > 140 |  |  |
| JumpSTART<sup>7</sup> | Walking? | Breathing<br>(Open Airway if<br>required) | If apnoeic assess for pulse. If<br>present give 5 rescue breaths. | Respiratory rate <15 or >45 | Palpable pulse? | Conscious level<br>assessment (AVPU) |  |
| Sheffield Paediatric Triage Tool (SPTT) | Catastrophic haemorrhage? | Walking? | Breathing<br>(Open Airway if required) | If apnoeic assess for pulse. If<br>present give 5 rescue<br>breaths. | Responds to voice? | Age appropriate<br>respiratory rate?* | Age appropriate heart<br>rate** |
| CareFlight<sup>14</sup> | Walking? | Obeys<br>commands? | Palpable radial pulse? OR<br>Breathes with open airway? | - | - | - | - |
| Major Incident Medical Management<br>and Support (MIMMS) Triage Sieve<sup>1</sup> | Walking? | Breathing<br>(Open Airway if<br>required) | Respiratory rate <10 or ≥30 | Heart Rate > 120 | - | - | - |
| Modified Physiological<br>Triage Tool 24 (MPTT-24)<sup>9,10,17</sup> | Catastrophic Haemorrhage? | Walking? | Breathing? Open Airway if<br>required. | Responds to voice? | Respiratory rate <12 or ≥24 | Heart rate ≥100 | - |
| Modified Simple Triage and Rapid<br>Treatment (MSTART)<sup>19</sup> | Walking? | Spontaneous<br>breathing | Respiratory rate >30 | Radial pulse absent | Obey commands | - | - |
| National Ambulance Service Medical<br>Directors (NASMeD) Triage Sieve<sup>18</sup> | Catastrophic haemorrhage | Are they injured? | Walking? | Breathing? Open airway<br>if required | Unconscious | Respiratory rate <10<br>or ≥30 | Pulse >120 or<br>capillary refill >2 sec |
| Rapid Assessment of<br>Mentation and Pulse<br>(RAMP)<sup>20</sup> | Casualty without signs of<br>obvious death | Casualty follows<br>commands | Radial pulse present? | - | - | - | - |

\*Respiratory Rate: <1: 30-40, 1-2: 25-25, 2-5: 25-30, 5-12: 20-25, >12: 15-20
\*\*Heart Rate: <1: 110-160, 1-2: 100-150, 2-5: 95-140, 5-12: 80-120, >12: 60-100.
\*\*\* MPTT-24 was updated in 2018 following consultation with NHS England to explicitly include the 'open airway' step as part of the breathing assessment.<sup>17</sup> This current version is currently in use in both UK military and civilian in-hospital practice (within the NHS Clinical Guidelines for Major Incidents).<sup>10</sup>

The primary outcome was the correct determination of Priority One status (requirement for life-saving intervention) in paediatric patients (defined as under 16 years) with subgroup analyses conducted for the age ranges <1yr, 1-2yrs, 2-5yrs, 5-12yrs, 12-16yrs, <12yrs, <16yrs. Secondary outcome measures included the prediction of major trauma (defined as Injury Severity Score (ISS) > 15) and mortality.

### Missing Data

A comparison was made between patients with complete data, and those missing pre-hospital physiological data, to explore for systematic differences in ISS, mortality and need for life-saving intervention. Performing a list-wise deletion on patients without complete data can introduce systematic errors, therefore multiple imputation was used on the first recorded physiological dataset in order to derive imputed data. Details of the modelling strategy are provided in supplementary file 3. In keeping with the main methods, a comparative analysis was then performed on the imputed data set.

### Statistical Analysis

Statistical analysis was performed to derive the sensitivity and specificity of the triage tools in detecting the outcomes of interest, and under triage (1-sensitivity) and over-triage (1-positive predictive value) were subsequently calculated. A chi-squared test was used to evaluate for statistical significance in categorical variables between included and excluded groups. Data distribution between the first-recorded physiology group and the imputed data group was compared using a Kolmogorov-Smirnov test. Python software (Version 3.7, Scotts Valley California, 2009) and R software (Version 3·6, R Core Team, New Zealand, 2000) were used for data processing and analysis.

### Patient and Public Involvement

Patients and/or the public were not involved in the design, or conduct, or reporting or dissemination plans of this research.

### Ethical Approval

TARN has ethical approval (Section 251) for research on anonymised data.

## Results

During the study period, 15133 patients aged under 16 years met TARN inclusion criteria, of which 4962 (32.8%) had complete pre-hospital physiological data and 8255 (54.5%) had complete first recorded physiological data (at scene and ED). A study flow diagram is shown in Figure 1 with age-group breakdown detailed in Table 2. Median age was 11.9 years (IQR 8.0-14.2), and the majority of patients were male (69.5%). Median ISS was 9 (IQR 9-17) with low overall mortality (1.1%). Blunt trauma predominated (95.4%), with motor vehicle collisions (49.6%) and low falls (23.9%), the leading mechanisms of injury. In total, 875 (17.6%) of patients with complete pre-hospital physiological data received at least one life-saving intervention and were considered Priority One, with advanced airway intervention (59.5%) predominating. Additional study characteristics are provided in Table 3.

**Figure 1:**
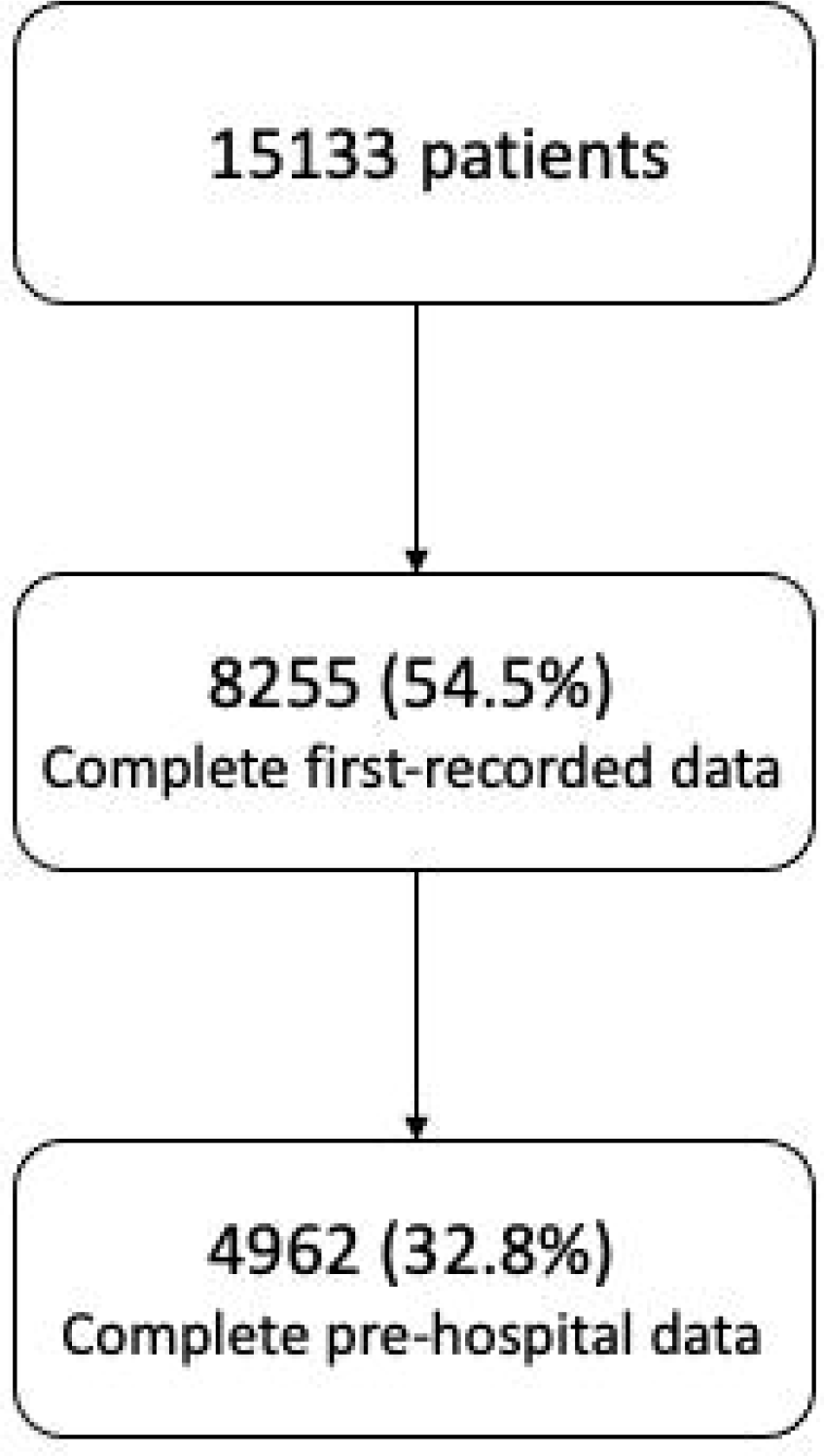
Study flow diagram.

**Table 2:** Age-group breakdown and data completeness.

| **Age** | **Under 1** | **1-2yrs** | **2-5yrs** | **5-12yrs** | **12-16yrs** |  |
| --- | --- | --- | --- | --- | --- | --- |
| Frequency (n,(%))* | 2072 (13.7%) | 1261 (8.3%) | 3045 (20.1%) | 4572 (30.21%) | 4183 (27.6%) | *N=15133 (100%)* |
| Complete pre-hospital physiology (n,(%)) | 145 (2.9%) | 59 (1.2%) | 445 (9.0%) | 1915 (38.6%) | 2398 (48.3%) | *N=4962 (32.8%)* |
| Complete first available physiology (n,(%))** | 508 (24.5%) | 260 (20.6%) | 1093 (35.9%) | 3095 (67.7%) | 3299 (78.9%) | *N=8255 (54.5%)* |
| Complete first available physiology (n,(%))*** | 1178 (56.7%) | 675 (53.5%) | 2044 (67.1%) | 3755 (82.1%) | 3631 (86.8%) | *N=11283 (74.6%)* |

\*Overall study population age < 16yrs, n=15133
\*\*Complete first available physiological data (ED and pre-hospital) physiological data
\*\*\*Complete first available physiological data (ED and pre-hospital) physiological using imputed data

**Table 3:** Characteristics of study population.

| Variable | Category/Stats | Complete Population<br>(n=15133) | Complete Pre-hospital Data<br>(n=4962, 32.8%) | Complete First Available Data<br>(n=8255, 54.5%) |
| --- | --- | --- | --- | --- |
| Gender | Male | 10294 (68.0%) | 3447 (69.5%) | 5682 (68.8%) |
|  | Female | 4839 (32.0%) | 1515 (30.5%) | 2573 (31.2%) |
| Injury Severity Score<br>(median (IQR)) |  | 9(9-16) | 9 (9-17) | 9 (9-16) |
| Age (years)<br>(median (IQR)) |  | 7 (2.3-12.5) | 11.9 (8-14.2) | 10.7 (5.6-13.8) |
| Outcome | Alive | 14764 (97.6%) | 4909 (98.9%) | 8188 (99.2%) |
|  | Dead | 369 (2.4%) | 53 (1.1%) | 67 (0.8%) |
| Mode of injury | Blunt | 14668 (96.9%) | 4733 (95.4%) | 7912 (95.8%) |
|  | Penetrating | 465 (3.1%) | 229 (4.6%) | 343 (4.2%) |
| Mechanism of injury (N (%)) |  |  |  |  |
|  | Fall less than 2m | 6014 (39.7%) | 1187 (23.9%) | 2374 (28.8%) |
|  | Vehicle incident/collision | 4721 (31.2%) | 2459 (49.6%) | 3616 (43.8%) |
|  | Fall more than 2m | 1514 (10.0%) | 645 (13.0%) | 1031 (12.5%) |
|  | Blow(s) | 1270 (8.4%) | 327 (6.6%) | 601 (7.3%) |
|  | Other | 1118 (7.4%) | 147 (3.0%) | 319 (3.9%) |
|  | Stabbing | 229 (1.5%) | 130 (2.6%) | 196 (2.4%) |
|  | Crush | 160 (1.1%) | 42 (0.9%) | 73 (0.9%) |
|  | Burn | 44 (0.3%) | 4 (0.1%) | 12 (0.2%) |
|  | Shooting | 42 (0.3%) | 16 (0.3%) | 24 (0.3%) |
|  | Blast | 21 (0.1%) | 5 (0.1%) | 9 (0.1%) |
| Priority One (N (%)) |  | 2820 (18.6%) | 1197 (24.1%) | 1813 (22.0%) |

The performance accuracy of all triage tools in the primary analysis is shown in Table 4a. The SPTT demonstrated the greatest sensitivity in determining Priority One status (92.2%, 95%CI 90.5-93.7%), followed by the MPTT-24 (80.8%, 95%CI 78.4-83.0%). The performance of the SPTT represents an absolute increase in sensitivity of 47.2-58.1% over other tools (PTT - 34.1% (95%CI 31.4-36.9%), JumpSTART – 45.0% (95%CI 42.1-47.8%)). This high sensitivity was correlated with low rates of under-triage (7.8% and 19.2% respectively for the SPTT and MPTT-24), but at the expense of higher rates of over-triage (75.0% and 70.2%) corresponding to specificities of 12.1% and 39.6% respectively.

**Table 4a:** Test characteristics with 95% Confidence Intervals – life-saving interventions.

| **Tool** | **Sensitivity** | **Specificity** | **Under triage** | **Over triage** |
| --- | --- | --- | --- | --- |
| SPTT | 92.2 (90.5, 93.7) | 12.1 (11.1, 13.2) | 7.8% | 75.0% |
| PTT | 34.1 (31.4, 36.9) | 85.8 (84.6, 86.9) | 65.9% | 56.7% |
| JUMPSTART | 45.0 (42.1, 47.8) | 92.1 (91.1, 92.9) | 55.1% | 35.7% |
| MPTT-24 | 80.8 (78.4, 83.0) | 39.6 (38.0, 41.2) | 19.2% | 70.2% |
| CARE_FLIGHT | 44.4 (41.5, 47.2) | 94.9 (94.1, 95.6) | 55.6% | 26.7% |
| MIMMS | 41.6 (38.8, 44.5) | 79.6 (78.3, 80.9) | 58.4% | 60.7% |
| NASMeD | 51.9 (49.0, 54.7) | 79.1 (77.8, 80.4) | 48.1% | 55.9% |
| RAMP | 43.8 (41.0, 46.6) | 94.9 (94.1, 95.6) | 56.2% | 26.8% |
| MSTART | 53.9 (51.0, 56.7) | 88.1 (87.0, 89.1) | 46.1% | 41.0% |

For detecting major trauma, the SPTT demonstrated the highest sensitivity (91.8%, 95%CI 90.3-93.1)) followed by the MPTT-24 (75.6%, 95%CI 73.5-77.7), with both PTT and JumpSTART demonstrating much lower sensitivity (28.8%, 95%CI 26.6-31.0; and 36.0%, 95%CI 33.7-38.4 respectively). For mortality, SPTT, MPTT-24 and JumpSTART had comparable sensitivities (83.0-88.7%), outperforming PTT (71.7%, 95%CI 57.7-83.2%). The full test characteristics for the secondary outcomes are provided Table 4b and 4c.

**Table 4b:** Test characteristics with 95% Confidence Intervals – ISS > 15.

| **Tool** | **Sensitivity** | **Specificity** | **Under triage** | **Over triage** |
| --- | --- | --- | --- | --- |
| SPTT | 91.8 (90.3-93.1) | 12.4 (11.3-13.6) | 8.2% | 66.4% |
| PTT | 28.8 (26.6-31.0) | 85.7 (84.5-86.9) | 71.2% | 50.7% |
| JUMPSTART | 36.0 (33.7-38.4) | 92.4 (91.4-93.3) | 64.0% | 30.5% |
| MPTT-24 | 75.6 (73.5-77.7) | 39.6 (38.0-41.3) | 24.4% | 62.3% |
| CARE_FLIGHT | 34.9 (32.6-37.3) | 95.2 (94.4-95.9) | 65.1% | 22.1% |
| MIMMS | 34.9 (32.6-37.3) | 79.0 (77.6-80.4) | 65.1% | 55.4% |
| NASMeD | 42.6 (40.2-45.1) | 78.5 (77.1-79.9) | 57.4% | 51.1% |
| RAMP | 34.5 (32.2-36.9) | 95.3 (94.5-96.0) | 65.5% | 22.1% |
| MSTART | 42.2 (39.8-44.7) | 87.7 (86.6-88.8) | 57.8% | 37.5% |

**Table 4c:** Test characteristics with 95% Confidence Intervals – Mortality.

| **Tool** | **Sensitivity** | **Specificity** | **Under triage** | **Over triage** |
| --- | --- | --- | --- | --- |
| SPTT | 86.8 (74.7-94.5) | 11.0 (10.2-12.0) | 13.2% | 92.2% |
| PTT | 71.7 (57.7-83.2) | 81.6 (80.5-82.6) | 28.3% | 96.0% |
| JUMPSTART | 88.7 (77.0-95.7) | 83.9 (82.9-84.9) | 11.3% | 94.4% |
| MPTT-24 | 83.0 (70.2-91.9) | 34.9 (33.5-36.2) | 17.0% | 98.6% |
| CARE_FLIGHT | 71.7 (57.7-83.2) | 86.0 (85.0-87.0) | 28.3% | 94.7% |
| MIMMS | 71.7 (57.7-83.2) | 75.0 (73.8-76.2) | 28.3% | 97.0% |
| NASMeD | 71.7 (57.7-83.2) | 72.1 (70.8-73.4) | 28.3% | 97.2% |
| RAMP | 69.8 (55.7-81.7) | 86.2 (85.2-87.1) | 30.2% | 94.8% |
| MSTART | 81.1 (68.0-90.6) | 78.6 (77.4-79.8) | 28.9% | 96.1% |

### Subgroup analysis by age category

The SPTT demonstrated a high sensitivity across all age groups (89.3% to 96.3%), with its lowest in the 5-12 year age group. The performance of PTT reduced as age increased, exhibiting the lowest sensitivity (20.8%, 95%CI 14.8-28.4%) for the 12-16 years age group. By contrast the performance of JumpSTART varied between the age groups and did not demonstrate a consistent trend. In the oldest age group (12-16years), the MPTT-24 demonstrated the greatest sensitivity (98.0%, 95%CI 93.8-99.5%) and apart from the SPTT, the remaining paediatric triage tools showed the worst performance across all age groups against which they were assessed. This subgroup analysis is provided in detail in supplementary table 2.

### Secondary analysis

First recorded physiological data (including pre-hospital and ED) were available for 8255 patients (54.5%). Within this cohort the median age was 10.4years (IQR 5.2-13.7) with males continuing to account for the majority of cases (68.7%). The outcome and median ISS remained unchanged (1.1% mortality, median ISS 9 (IQR 9-17)) to the primary analysis. A comparable proportion (16.7 vs 17.6%) received at least one life-saving intervention, with advanced airway intervention again predominating (60.1%).

Tool performance was largely similar to the primary analysis, with the SPTT exhibiting the highest sensitivity (90.0% (95%CI 88.5-91.2%), followed by the MPTT-24 (81.2% (95%CI 79.5-82.9%)). The full secondary analysis test characteristics are provided in supplementary table 3.

### Missing Data

A significant proportion of patients had incomplete pre-hospital physiological data (n=10171, 67.2%). Those with missing data were significantly younger (median age 3.9 vs 11.9, p<0.0001). Whilst no difference was observed in median ISS, the outcome differed between the two groups with a higher mortality in those excluded (3.1% vs 1.1%, p<0.0001). Additionally, the leading mechanisms of injury were ‘reversed’ between the complete (motor vehicle collision 49.6%, low falls 23.9%) and incomplete data groups (motor vehicle collision 22.2%, low falls 47.5%).

Performance characteristics were unchanged following multiple imputation to account for missing data. The full test characteristics are provided in supplementary table 3b.

## Discussion

Using a large civilian trauma registry, we have validated the performance accuracy of the novel SPTT and compared it with existing adult and paediatric triage tools in identifying the need for life-saving interventions in a UK paediatric population. The SPTT (a specific paediatric tool) and the MPTT-24 (an adult triage tool) are the most accurate at predicting need for life-saving intervention, major injury and mortality in the paediatric population.

While there has been much recent focus on defining the optimal major incident triage tool for adult patients, this has not yet been the case for children. Few bespoke paediatric triage tools have been derived and there is minimal data from external validations to support the use of one over another on the basis of performance characteristics.^2,15,21-23^ Whilst previous studies have compared triage tool performance using either mortality or ISS, neither of these are likely to extrapolate well to the acuity of the patient whilst in the pre-hospital setting.^2-4^ In a major incident, the purpose of triage is to prioritise those patients who may benefit from life-saving interventions.^2,5,24^ This outcome measure should therefore form the primary outcome of interest in the assessment of any such tool. The performance accuracy of the triage tools assessed in this study was further delineated using subgroup analysis by age range, and extended to the secondary outcomes of major trauma and mortality.

The first paediatric tool, PTT^6^, was developed to prevent over-triage when the adult Triage Sieve was used on paediatric patients. In a previous comparative analysis, PTT showed high specificity (>98%) for identifying patients who either required life-saving interventions, or who had sustained major trauma (ISS > 15). However, despite good specificity, the tool demonstrated poor sensitivity for both outcomes, corresponding to under-triage rates in excess of 58%.^2^ In the same study, JumpSTART, whilst also performing with high specificity (>97%), demonstrated less than 5% sensitivity for both outcomes, correlating with under triage rates in excess of 95%.^2^ A further study used trauma registry data to assess performance accuracy of triage tools in predicting mortality and major trauma, and reported similar sensitivity levels for the PTT but considerably lower specificity (66.0% and 66.5% respectively). By contrast, JumpSTART demonstrated better performance.^21^ With life-saving intervention being our primary outcome measure, our study aligns more closely to the former study, and whilst we report a comparable performance in sensitivity for the PTT (37.3% vs 41.5%), the JumpSTART method exhibited improved performance against our dataset (45.3% vs 0.8%).^2^

The key principles of triage are that it should be rapid, reliable and reproducible, irrespective of the provider performing it.^1^ The reliability of the triage tool is the assessment of its performance; key to which is identifying those in need of life-saving interventions and minimising under-triage (the misclassification of patients as not-needing a life-saving intervention). In an ideal setting, the triage tool used would minimise both under and over-triage, but the reality is that increasing sensitivity often corresponds to decreasing specificity necessitating a decision over their importance. An additional factor, when assessing triage methods, lies with their application simplicity; an overly complex triage system with good performance may not be practical, particularly within pre-hospital settings. Whilst the SPTT demonstrated the highest overall sensitivity in our study, the inclusion of five age categories with different physiological variables is unlikely to make it a practical option for use in either pre-hospital settings or in surge situations within hospital settings.

Specific paediatric tools also differ in the ages in which the tools are recommended (JumpSTART^7^ and the PTT^6^ differ in their approach, recommending cut-offs at 8 and 12 years respectively). For example, the subgroup analysis of the 12-16 year age group in this study demonstrates directly comparable median physiological parameters to that observed in a previous adult major incident triage study, which would support the approach taken by the PTT.^6,25^

One potential solution is the application of a single tool across all age ranges, covering adult and paediatric patients. This may need to involve a compromise between optimal tool performance, practicality and ease of use. The adoption of single physiological thresholds, such as those used in the MPTT-24^9^, represents a more simplistic option and will convey additional benefit from the perspective of familiarity and training. However, as observed within the MPTT-24 performance analysis, whilst this reduces under-triage, it is associated with increased over-triage, albeit comparable to that tolerated within the adult setting.

## Limitations

A key limitation of our work lies with the use of a retrospective trauma database. Firstly, the mechanism of injury encountered on the database (RTCs and low falls) is unlikely to accurately represent the injury pattern encountered following a major incident in its totality. Ideally, any analysis of triage tools should be performed in the environment in which they are designed to function but owing to the unpredictable nature of major incidents this is both impractical and also largely unethical as well as unrealistic given the frequency of paediatric major incidents. As a result, trauma databases are frequently used as a surrogate, allowing for the analysis of a large number of seriously injured patients.^9,14,15^ A further limitation is the presence of inclusion criteria for entry into the TARN database; whilst only a minority of patients (17.6%) received a life-saving intervention in our analysis, the inclusion criteria will likely skew the study population towards those with more severe injuries. Therefore, it would be anticipated that the frequency of patients not receiving a life-saving intervention in the population will be higher than observed in this study.

We also acknowledge that the exclusion of patients with incomplete physiological data is an additional limitation of our study, with only 32.8% having complete pre-hospital data with which to perform the primary analysis. Whilst median ISS is comparable between the included and excluded groups, we did observe a difference in median age and outcome, with those patients in the excluded group being younger and having a higher mortality. In an attempt to mitigate for the missing pre-hospital data, additional analyses were conducted using first recorded physiological data (including ED data) and also on an imputed dataset. However, even this is imperfect as physiology may have changed by arrival to ED in response to any interventions performed in the pre-hospital setting. Performance characteristics of the triage tools were unchanged in these further analyses.

## Conclusion

In this comparative analysis of paediatric triage tools, the SPTT and MPTT-24 perform better than existing paediatric triage tools for identifying those patients requiring life-saving intervention. This may necessitate a change in recommended practice. Further work is required to determine the optimum method of paediatric major incident triage, but consideration should be given to simplifying major incident triage by the use of one generic tool for adults and children.

## Data Availability

De-identified patient data utilised for this study are proprietary to the Trauma Audit and Research Network, University of Manchester and may be requested directly from TARN

## Author contributions

JV, ML, JES designed the study. JV, SC, NM, YX verified the underlying data. JV and SC conducted the analysis. All authors contributed to data interpretation. JV wrote the initial draft of the manuscript. All authors contributed with critical revisions to the manuscript. JV takes responsibility for the manuscript as a whole.

## Statements

## Ethical Approval

TARN has ethical approval (Section 251) for research on anonymised data.

## Clinical Trial Registration

Not applicable.

## Acknowledgements

The authors thank Prof Fiona Lecky (research director) and Antoinette Edwards (Chief Executive Officer) at Trauma Audit Research Network (TARN) for facilitating access to the TARN database.

