## Supplementary Files for "Comparative Analysis of Major Incident Triage Tools in Children – a UK population-based analysis"

Supplementary File 1: TARN inclusion criteria

Supplementary File 2: Assumptions for characterisation by triage tools.

A number of assumptions were made in order to facilitate the retrospective application of triage tools. In keeping with previously published studies, patients with a GCS ≤ 8 were categorised as unconscious, and those with a GCS < 13 were deemed to be unresponsive to voice.^9,13^

Patients with a GCS Motor component less than 6 were deemed to not be able to obey commands. A systolic blood pressure of 60mmHg was used as a surrogate for the presence of a palpable pulse.

Owing to the nature of the TARN registry, presence of catastrophic haemorrhage is not a collated variable and therefore was unable to be applied by triage tools including this assessment.

Patients who were recorded on TARN as having a ‘supported’ or ‘obstructed’ airway at scene were regarded as having received an airway opening manoeuvre.

Where a weight calculation was required (either for classification of triage category i.e. by the PTT^6^ or for determination of fluid requirement as per Lerner criteria with APLS modification^12,16^), standard APLS formula were used based on the age of the patient.

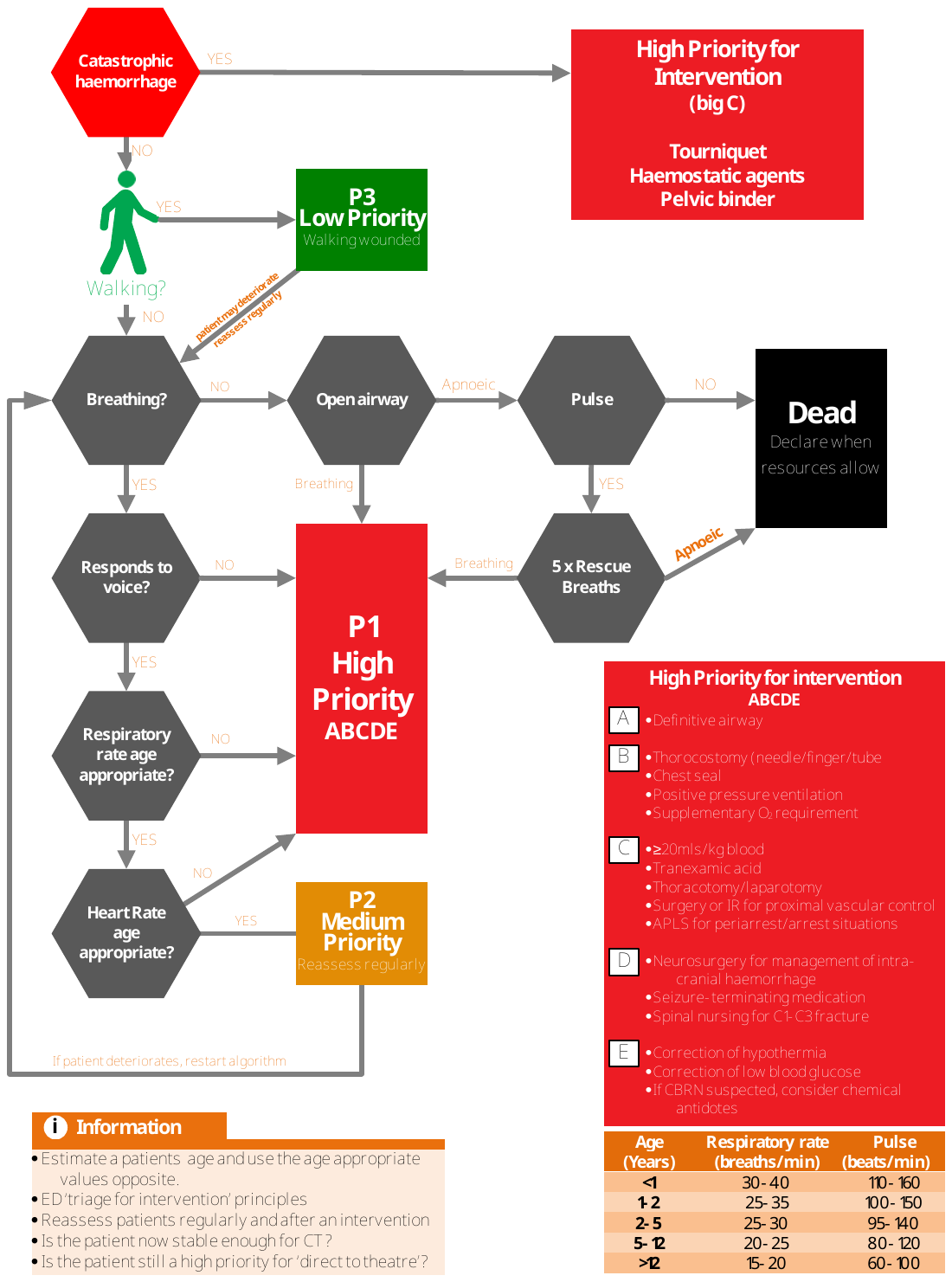

**Supplementary Figure 1: Sheffield Paediatric Triage Tool (SPTT)**

**Supplementary Table 1: Lerner criteria defining life-saving interventions^16^**

Limb-conserving surgery performed within 4 hours of arrival at hospital on a limb that was found to be pulseless distal to the injury prior to surgery

Neurologic, vascular, or haemorrhage-controlling surgery to the head, neck or torso performed within 4 hours of arrival to hospital.

Chest tube placed within 2 hours of arrival at hospital

Escharotomy performed on a patient with burns within 2 hours of arrival at a hospital

An advanced airway intervention (e.g. intubation, LMA, surgical airway) performed in the pre-hospital setting

IV vasopressors administered within 2 hours of arrival at hospital or within 4 hours of arrival at hospital

Patient who required EMS initiation of CPR (i.e. had a cardiac arrest) during transport, in the ED, or within 4 hours of arrival at a hospital

Arrived in the ED with uncontrolled haemorrhage

**Supplementary Table 2: Test characteristics with 95% Confidence Intervals for subgroup analysis.**

| **Under 1yrs** | **1-2yrs** |
| --- | --- |
| 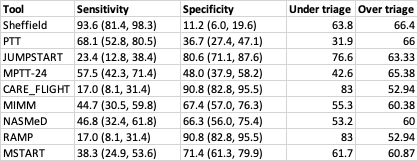 | 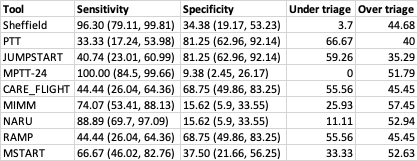 |
| **2-5yrs** | **5-12yrs** |
| 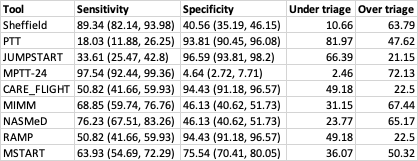 | 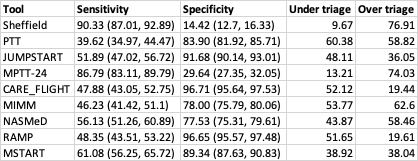 |
| **12-16yrs** | |
| 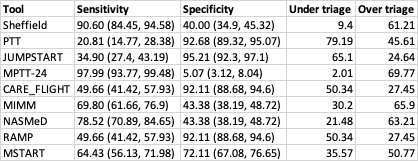 | |

**Supplementary Table 3a: Test characteristics with 95% Confidence Intervals for secondary analysis (first recorded physiology).**

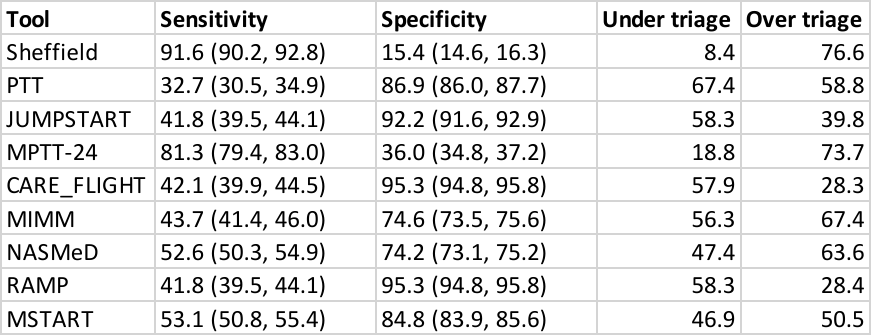

**Supplementary Table 3b: Test characteristics with 95% Confidence Intervals for imputed dataset**

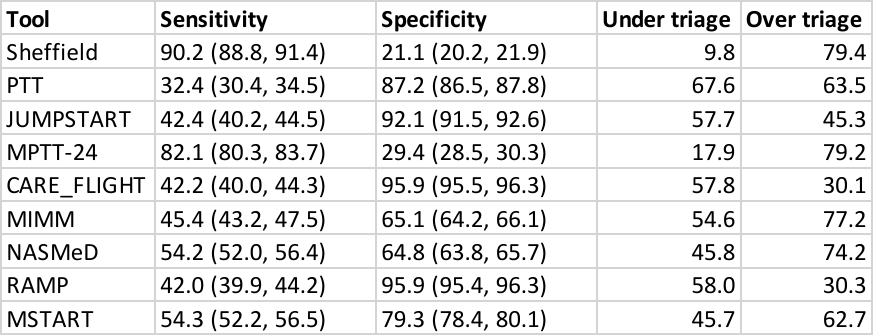

Supplementary File 3: Multiple imputation details

First available physiological data was used for the multiple imputation process. Pre-processing was conducted in order to remove incorrect data from the dataset. Subsequently patients missing more than two physiological variables (Respiratory Rate, Heart Rate, Systolic Blood Pressure, GCS) were excluded.

An iterative regression model (Multiple imputation using Chained Equations – MICE) was used to derive the imputed data. Patients data including age, gender, injury type and mechanism, ISS, AIS, physiology, and LSI were used in imputed model. Patients whose GCS total is not equal to the sum of the three components were removed. 5 imputed data sets were generated and the data set with the smallest number of incorrect GCS total was chosen for further analysis. Following imputation this resulted in 11,952 patients with which to perform the comparative analysis.
